# The pioneering gut microbiome acquired via different delivery modes in neonates shapes distinct immune and metabolic environments

**DOI:** 10.1101/2025.09.30.25337005

**Authors:** Sivaranjani Namasivayam, Hector N Romero-Soto, Mickayla Bacorn, P. Juliana Perez-Chaparro, Andrew S Burns, Shreni Mistry, Nathan T. Brandes, Audrey Chong, Gwendolyn Cooper, Ian S. LaCroix, Phoebe LaPoint, Keona Banks, Qing Chen, Anal Patel, Noel T. Mueller, Maria Gloria Dominguez-Bello, George L. Maxwell, Shira Levy, Benjamin Schwarz, Yasmine Belkaid, Suchitra K. Hourigan

## Abstract

**Background:** Cesarean section (CS) delivery is associated with an increased risk of inflammatory diseases, hypothesized to be driven by differences in the microbiome acquired at birth compared to vaginally delivered (VD) infants. How delivery mode associated differences in initial colonizers directly modulate early life immune education and metabolic development is poorly understood.

**Objective:** First, to investigate how differences in pioneering colonizers associated with delivery mode directly modulate early life immune education and metabolic programming. Second, to examine the effect of “vaginal seeding”, an intervention aimed to restore the microbiome of CS infants to a VD state.

**Design:** Germ-free mice were colonized with transitional stool from VD, CS or CS-delivered and vaginally seeded neonates. Immune cell populations, serum immunoglobulin levels, and fecal microbiome and metabolome profiles were analyzed.

**Results:** Mice colonized with stool from VD neonates displayed increased numbers of myeloid cells at barrier tissues, whereas CS microbiome colonized mice exhibited decreased Th1/Th2 ratios and increased serum IgE levels. Key amino-acid pathways including tryptophan metabolism, riboflavin co-enzymes and carbohydrate metabolites were significantly enriched in the murine VD fecal metabolome and correlate with the increased abundance of *Escherichia* typically observed in the VD microbiome. Mice colonized with stool from CS neonates who received vaginal seeding, resulted in increased regulatory T cells and serum IgA in mice, suggesting potential benefits of vaginal seeding.

**Conclusion:** Collectively, our studies demonstrate the ability of pioneering colonizers to set the immune and metabolic tone that could have long-lasting effects and provide avenues for microbiome-mediated interventions.

## INTRODUCTION

The early life microbiome plays a pivotal role in the education of the developing neonatal immune system, establishment of metabolic homeostasis and immunometabolic programming^1,2^. The nature and quality of the host-microbiome relationship need to be well-orchestrated during this critical “window of opportunity”^3,4^. Germ-free murine models have demonstrated that the absence of microbes leads to an allergic immune tone and reduced regulatory T cell function among other defects in the immune landscape, several of which can only be rescued if the mice are colonized with a complete or defined microbial community only within the first few weeks of life^5–7^. Similarly, disruption of the microbiome in early life via antibiotics leads to obesity later in life^8,9^. Importantly, numerous clinical studies have demonstrated that lifestyle and environmental factors impact early life host-microbiome interactions which in turn influence the trajectory of host health^10^.

Rapid microbial colonization and expansion at the mucosal sites occurs at birth^11–13^. As such, epidemiological studies have identified delivery mode as a key determinant in shaping the developing microbiome, immune and metabolomic systems^14–18^. Indeed, cesarean section (CS) birth has been associated with increased risk of inflammatory and metabolic diseases such as obesity and atopy, including asthma, and inflammatory bowel disease, that is attributed to, at least in part, different microbes acquired during CS delivery compared to a vaginally delivered (VD) infant^19–26^. Neonates born via CS are seeded with the mother’s skin and environmental microbes that is characterized by *Enterococcus*, *Klebsiella*, *Staphylococcus*, *Corynebacterium* and *Propionibacterium* spp. in the stool within the first week of life^27,28^. In contrast, a VD neonate is colonized during passage through the vaginal canal with maternal perineal-vaginal microbes, with *Bifidobacterium*, *Escherichia*, *Bacteroides* and *Lactobacillus* being the predominant taxa in early VD neonatal stool^27–29^. Variations in the composition and maturation of the microbiome between the two delivery modes can persist for over a year, with the CS microbiome associated with delayed maturity^14,30–32^.

Vaginal seeding (VS), the wiping down of the CS delivered neonate with the mother’s own vaginal fluid immediately following birth is being investigated as a microbiome-mediated clinical intervention to transfer the mother’s vaginal microbiota to the CS infant to compensate for bypassing the vaginal canal^33–36^. This method has been demonstrated to allow at least partial engraftment of the maternal vaginal microbiome at multiple body sites of the CS delivered neonate^37,38^. More importantly, a recent clinical trial linked VS to potential benefits in neurodevelopment in CS delivered infants^39^. Furthermore, using a human-to-murine fecal transplant model we demonstrated the ability of the microbiota from CS delivered and vaginally seeded neonates to reduce intraabdominal adiposity in comparison to microbes from CS delivered neonates without VS^40^.

While several studies have demonstrated the significance of the mother’s vaginally acquired microbiome for the infant’s health, how the microbiomes acquired by different delivery modes directly modulate the trajectory of the infant’s health remains poorly understood. Specifically, the role of the pioneering colonizers in setting the baseline immune tone is unknown. Therefore, employing a humanized microbiome murine model, we first examined how VD and CS acquired pioneering microbiomes differentially modulate the immune and metabolic environments and second, investigated whether and how VS modifies these landscapes.

## RESULTS

### Colonization of germ-free mice by distinct human neonatal microbiomes acquired at birth

Transitional stool from neonates in the first few days of life, displays significant differences in the microbial composition by delivery mode while still being reflective of the initial bacterial colonizers of the infant gut, and hence was used in this study^28^. Transitional stool from neonates that were delivered vaginally (VD), via C-section (CS), or C-section delivered and vaginally seeded (CS+VS) were orally inoculated into independent groups of mice at the time of weaning of the mice from their mothers (Figure 1A, Supplemental Table 1 for clinical and demographic characteristics of infant cohort). Microbiota bacterial composition in the small intestine, colon and fecal pellets of colonized mice was assessed three weeks post-inoculation via *16s rRNA* gene sequencing.

**Figure 1.**
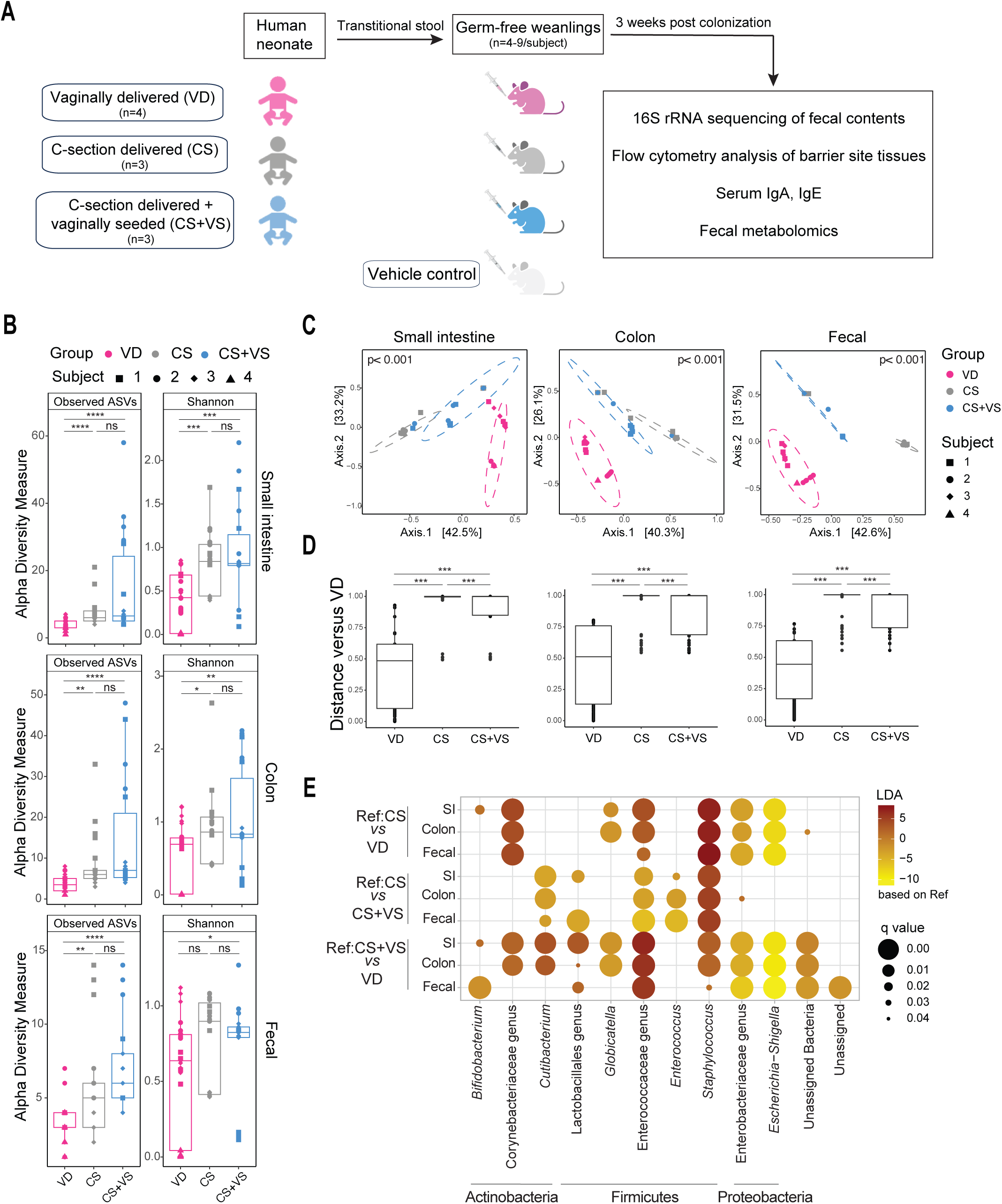
Composition of gut microbiomes in mice colonized with stool from neonates born via different delivery modes. **A**. Schematic of experimental design. Stool samples from vaginally delivered (VD), cesarean section (CS) delivered or CS-delivered and vaginally seeded (CS+VS) neonates were orally inoculated into germ-free mice at weaning. An additional group of germ-free mice received PBS as vehicle control. Three weeks post-inoculation fecal, serum and tissues samples were collected for downstream analysis. Microbiome profiling was performed on fecal samples collected from small intestinal and colon flushes, as well as fecal pellets, using *16S rRNA* gene sequencing. **B.** Alpha diversity estimates of the microbiota from the small intestine, colon and fecal pellets were calculated using Observed ASVs and Shannon index. Each dot denotes a mouse with the shape and color representing a human donor subject and experimental group used to inoculate the mouse respectively, as shown in the key. Statistical significance was calculated using Kruskal-Wallis test (ns, p > 0.05; * p < 0.05; ** p < 0.01; ***p < 0.001; ****p < 0.0001) **C.** Principal coordinate analysis (PCoA) plots of Bray Curtis dissimilarity metrics colored by inoculation group. Statistical significance was assessed using Adonis with 999 permutations. **D**. Beta-diversity distances within and between groups using VD group as reference are plotted and statistical significance was calculated using PERMANOVA (***p <0.001) **E.** Differentially abundant taxa were identified using ANCOMBC for each pairwise comparison, as indicated along the x-asis and displayed as a dotplot. Genera that were signficantly different (q < 0.05) are grouped by phyla along the y-axis. ASVs not classified with a specific genus are indicated as a genus under the highest taxonmic classification reported or as unclassified. Colors represent differential abundance relative to the reference group (Ref) and size of the circles indicate signficance values following multiple-testing as shown in the key.

We observed significantly lower alpha diversity, assessed via observed ASVs and Shannon index, in the microbiome of mice that received stool of VD neonates (VD mice) in comparison to mice that were colonized with stool from CS (CS mice) or CS+VS neonates (CS+VS mice) (Figure 1B). Alpha diversity measures were not significantly different between CS and CS+VS mice. However, when assessing beta-diversity based on Bray Curtis dissimilarity distance matrix, microbiota community structure was significantly different between the three groups of mice at all three intestinal sites (Figure 1C, 1D).

We next compared the composition of the bacterial microbiota across groups to identify significantly altered taxa. Assessment at the family level indicated distinct microbial taxonomic profiles in VD, CS and CS+VS groups (Figure S1A). The intestinal microbiota in VD mice, was characterized by significant enrichment in an unclassified genus from the Enterobacteriaceae family and *Escherichia/Shigella* (Figure 1E, Figure S1B). CS mice displayed increased abundance of *Staphylococcus*, and unclassified genera of families Corynebacteriaceae and Enterococcaeceae, when compared to VD mice (Figure 1E, Figure S1B). Interestingly, while *Staphylococcus* was significantly enriched in CS+VS mice in comparison to VD mice, this genus was also increased in CS mice relative to CS+VS mice, suggesting that VS may help reduce the abundance of *Staphylococcus* typically seen in the CS microbiome (Figure 1E, Figure S1B). *Cutibacterium and* unclassified genera of order Lactobacillales and family Enterococcaceae were enriched in CS+VS mice intestinal microbiota in comparison to the microbiome of both VD and CS mice with a higher effect size observed when compared to VD mice (Figure 1E, Figure S1B). *Enterococcus* was increased in CS+VS mice only in comparison to the CS mice microbiome. Interestingly, *Bifidobacterium* is enriched in both CS and CS+VS mice in comparison to VD mice but not uniformly at all intestinal sites (Figure 1E, Figure S1B). It should be noted that interindividual variability among subjects does exist especially within the CS+VS group. However, together, these data indicate successful colonization of germ-free mice with distinct microbial communities typically observed in VD and CS neonates, with the VS intervention resulting in defined alterations in comparison to the CS microbiome.

### Microbial communities modulate the myeloid immune landscape at barrier sites of colonized mice

We next assessed how the distinct microbes acquired via different delivery modes modulate the immune landscape at major barrier sites of gut, lung and skin of colonized germ-free mice via flow cytometry. In the small intestine lamina propria (siLP), among quantified granulocytes, neutrophils were elevated in CS+VS mice compared to both VD and CS animals and no differences were evident in eosinophils (Figure 2A). We observed no significant differences in Ly6C+ monocytes or the macrophage population. However, in terms of the dendritic cell subsets, CD103^+^CD11b^+^ DCs and CD11b^+^ DCs were significantly lower in CS+VS (with only CD11b^+^ DCs significantly lower in CS) in comparison to the VD group (Figure 2A). Interestingly, we observed more profound differences in the myeloid cell population in the lung and the skin between the groups.

**Figure 2.**
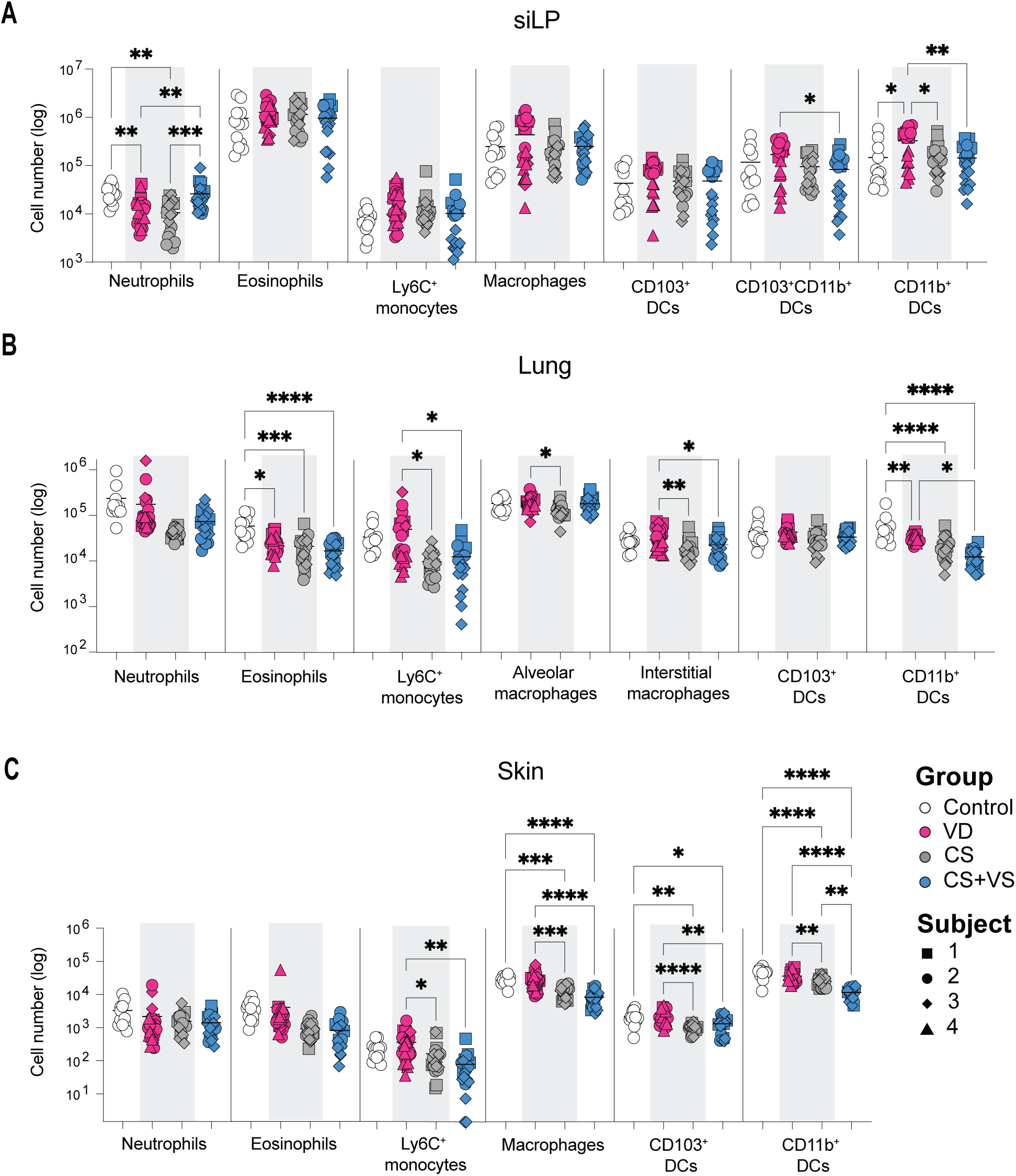
Influence of different neonatal microbiomes acquired at birth in shaping the myeloid compartment at various barrier tissues in colonized germ-free mice. **A-C.** Quanitification of major myeloid cell populations in the siLP (A), lung (B) and skin (C). Each dot denotes a mouse with the shape and color representing a human donor subject and experimental group used to inoculate the mouse respectively, as shown in the key in C. Control group represents germ-free mice inoculated with PBS vehicle control. The gating strategy is as shown in Figure S2A. Statistical significance was assessed by one-way ANOVA with Tukey’s multiple comparison test and only statistically signficant comparisons are shown (not shown, p > 0.05; * p < 0.05; ** p < 0.01; ***p < 0.001; ****p < 0.0001)

While we did not observe significant differences in the neutrophil numbers in either the lungs or skin, eosinophils in the lungs of the colonized mice were decreased in comparison to germ-free controls (Figure 2B-C). In contrast to the siLP, Ly6C^+^ monocytes and macrophages were significantly higher in the lung and skin of VD mice when compared to CS and CS+VS mice. Alveolar macrophages in the lung were decreased only in the CS group (Figure 2B). In line with observations in the siLP, VD mice displayed increased numbers of CD11b^+^ DCs in the lung and skin whereas, tolerogenic CD103^+^ DCs were elevated only in the skin of VD mice (Figure 2B-C). Overall, the myeloid compartment of VD microbiome colonized mice displays increased cell numbers suggesting potentially increased immune activation, that is only minimally restored by VS of the CS neonates (e.g. increased neutrophils in the siLP of CS+VS mice).

### Vaginal seeding partially restores the lymphoid compartment at barrier tissues to a VD state

We next compared the lymphoid cell population in the colonized mice at the same barrier tissues in addition to a secondary lymphoid organ, the gut draining mesenteric lymph node (LN). We quantified the numbers and frequency of CD4^+^ and ψ8 T cell subsets, number of CD8^+^ T cells, NK cells and innate lymphoid cells (ILCs) in the siLP, lung and LN. Only numbers of CD4^+^, CD8^+^, ψ8 T and NK cells were examined in the skin as too few cells were present to further identify phenotypic subsets. When comparing colonized animals, we observed increased CD4^+^ T, CD8^+^ T and NK cells in the siLP, elevated NK cells in the lungs and γδ T cells in the skin of VD mice (Figure 3A-C). γδ T cells in the siLP alone were increased in CS+VS mice relative to CS mice. No significant differences in these cell types or ILCs were observed in the other tissues across colonized mice (Figures 3A-C, S3A).

**Figure 3.**
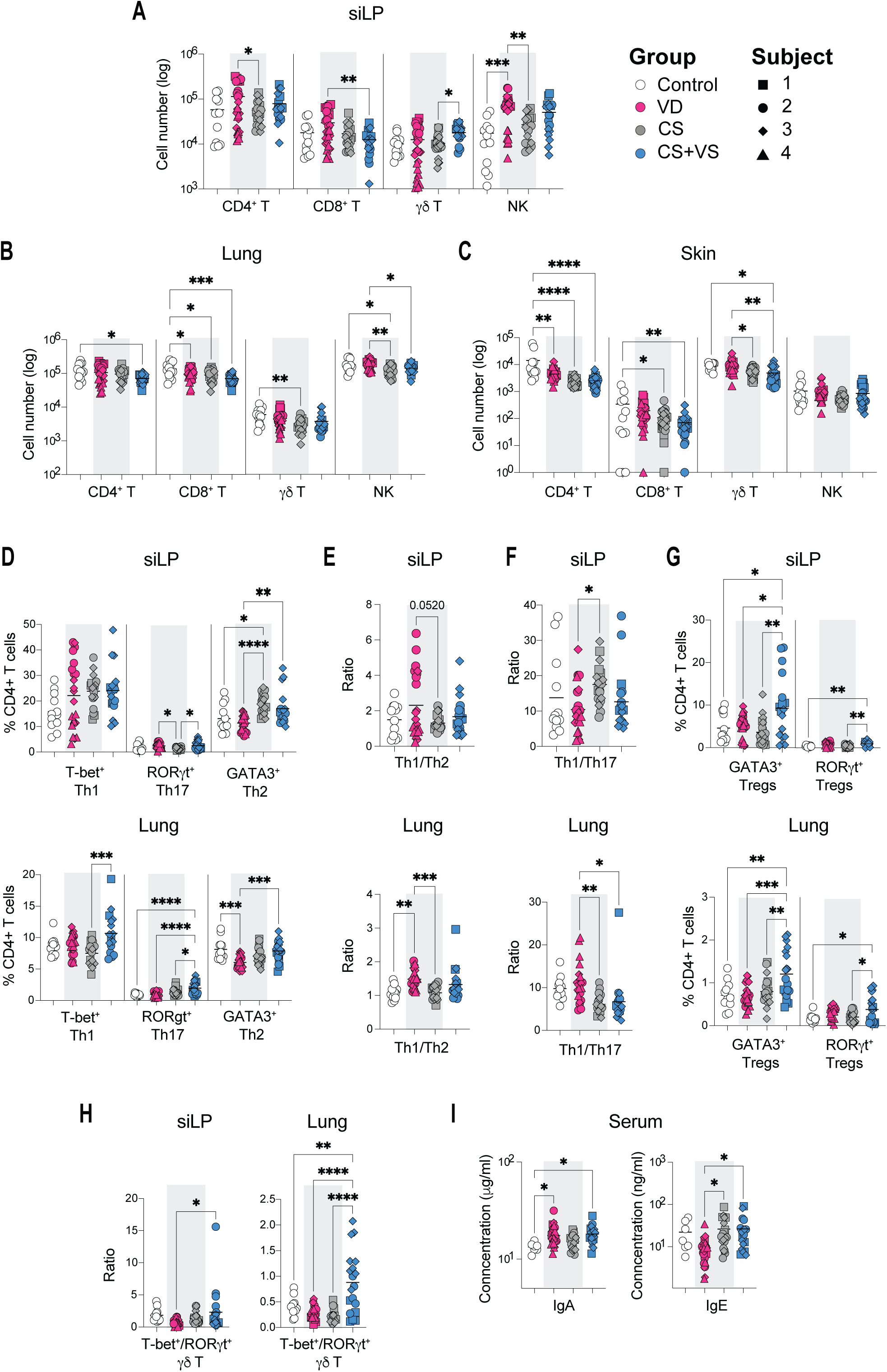
Comparison of the lymphoid compartment at barrier tissues in neonatal stool colonized mice. **A-C.** Cell numbers of major lymphocyte cell types in the siLP (A), lung (B) and skin (C). **D.** Frequency of CD4^+^ T cell subsets in siLP and lung. **E.** Ratio of Th1/Th2 cells. **F.** Ratio of Th1/Th17 cells. **G**. Frequency of Treg subsets. **H.** Ratio of Tbet^+^/RORγt^+^ γδ T cells. **I.** Concentration of IgA and IgE in serum. The gating strategy is as shown in Figure S2B. Each dot represents a mouse and is coded as shown in the key. Statistical signifcance was assessed by one-way ANOVA with Tukey’s multiple comparison test and only statistically signficant comparisons are shown (not shown, p > 0.05; * p < 0.05; ** p < 0.01; ***p < 0.001; ****p < 0.0001)

Among the conventional effector CD4^+^ T cell subsets, CS mice were characterized with increased frequency of GATA-3^+^ Th2 cells and decreased frequency of RORγt^+^ Th17 in the siLP with similar trends in the LN and lungs (Figure 3D, Figure S3C-D, S4B-C). This resulted in a decreased Th1/Th2 ratio, indicative of an allergic environment, in siLP, LN and lungs of CS mice in comparison to VD mice (Figure 3E, Figure S4D). This ratio was moderately increased in CS+VS animals and was similar to that observed in the lungs of VD mice (Figure 3E). In contrast, CS mice displayed increased Th1/Th17 ratio in the siLP and LN, while this ratio was decreased in the lungs of these animals (Figure 3F, Figure S4D). CS+VS mice showed decreased Th1/Th17 ratio in all tissues, overall, suggesting a differential modulation of these T cell subsets by the different microbiota communities. In terms of cytokine production capacity, in the LN, we observed increased IFNγ^+^ CD4^+^ and IFNγ^+^ CD8^+^ T cells in CS and CS+VS mice, whereas IL22^+^ CD4^+^ T cells were increased in VD mice (Figure S4E-F). With regards to the regulatory T cells (Tregs) CS+VS mice displayed elevated levels of GATA-3^+^ and RORγ^+^ Tregs in the siLP and lung (Figure 3G). Importantly, microbiota dependent RORγt^+^ Tregs were significantly higher in CS+VS mice in comparison to CS animals (Figures 3G).

In the case of γδ T cells, Tbet^+^ γδ T in the lungs and IFNγ^+^ γδ T cells in the LN were significantly increased in CS+VS mice, while RORγt^+^ γδ T were decreased in both the siLP and lungs of these animals (Figure S3E-F, Figure S4G-H). In contrast, RORγt^+^ γδ T and IL17A^+^ γδ T cells were elevated in VD mice. These differences resulted in significantly increased Tbet^+^/RORγt^+^ γδ T ratio in the CS+VS animals (Figure 3H, Figure S4I).

Finally, we quantified the serum concentrations of IgA and IgE, immunoglobins known to be modulated by the microbiome in early life^5^. We observed that IgA was significantly increased in the serum of VD and CS+VS but not CS mice when compared to germ-free control animals (Figure 3I). However, only VD mice had lower concentrations of IgE when compared to both CS and CS+VS animals. Taken together, these findings demonstrate that microbiomes derived from CS neonates display a Th2 skew with a decreased Th17 and regulatory T cell environment in colonized mice, that is partially restored in mice colonized with microbiota from CS+VS neonates.

### Delivery mode acquired microbiota distinctly shape the fecal metabolome of colonized mice

Next, we examined how the different communities of delivery mode acquired microbes shape the intestinal metabolic balance in mice though mass spectrometry based targeted metabolomics on fecal pellets collected three weeks post-inoculation. As expected, the metabolic profiles of colonized mice are distinct from that of germ-free mice, with germ-free mice displaying increased levels of nucleobases and nucleosides (Figure S5). More importantly, within the colonized groups, we observed that metabolic profiles of mice colonized with VD microbiome were distinct (Dim 1: 32.7% variance) from that of CS and CS+VS microbiota colonized mice, as determined through principal component analysis (PCA) (Figure 4A). The size of the metabolic difference associated with the VD microbiome was large with 88 metabolites in the VD versus CS and 116 metabolites in VD versus CS+VS comparisons, of the 196 targeted metabolites, found to be significantly (adjusted p < 0.05) altered with a log_2_ fold change of at least 0.5 (Figure 4B, Figure S5A). Specifically, dominant differences across the colonized groups included metabolites with known roles in gut homeostasis and immune function, including tryptophan, histamine, acetylcholine, arginine/amine metabolites, redox cofactor and central energy metabolites (Figure 4B, Figures S5A, S6, S7).

**Figure 4.**
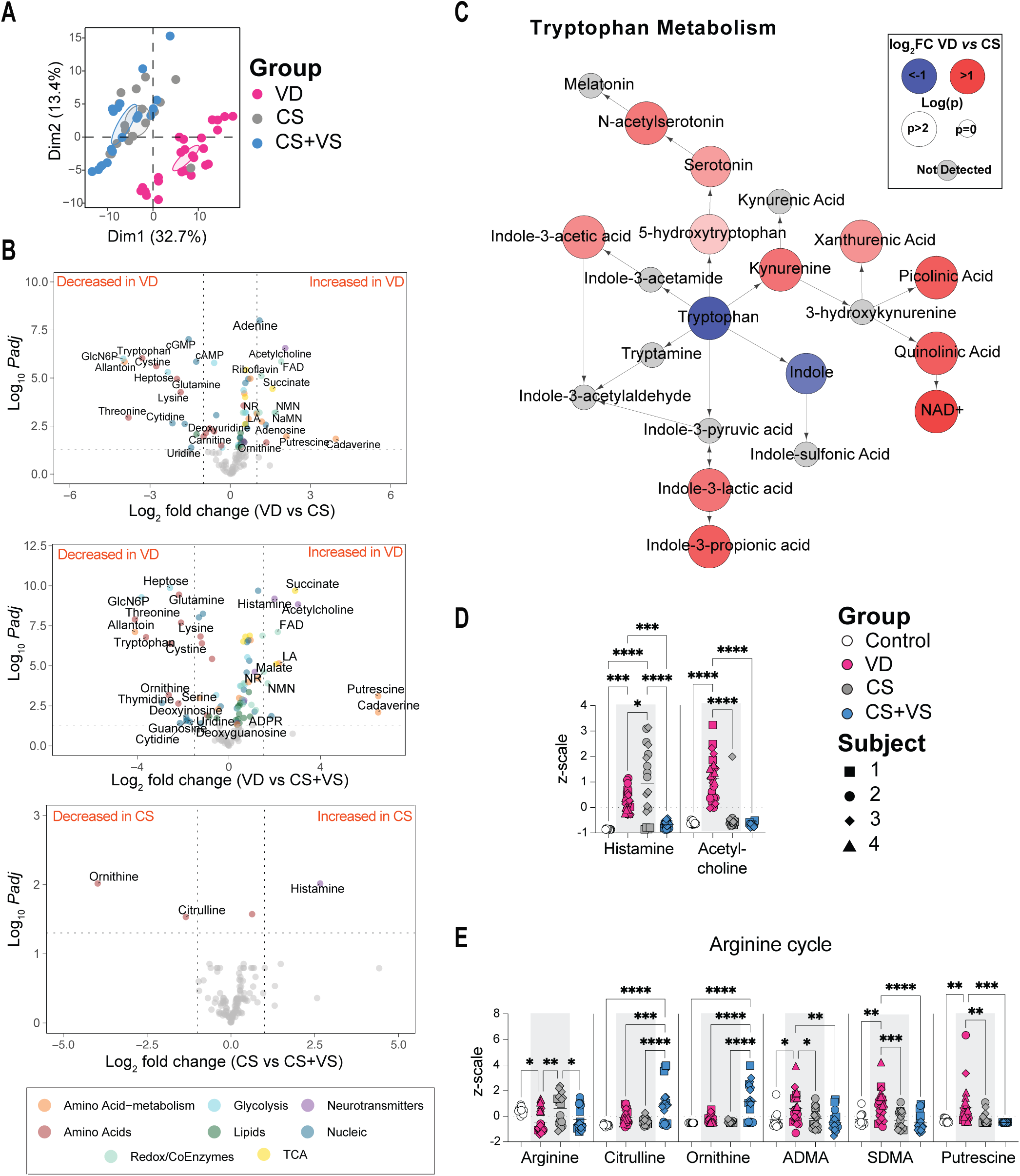
Comparison of fecal metabolites of mice colonized with neonatal stool. **A.** PCA plot of fecal metabolite signals identified from mice colonized with VD, CS, or CS+VS neonate stool **B.** Volcano plots show metabolites that were differentially abundant in pairwise comparisons of groups as indicated. Statistical significance was assessed using Student’s t-test and metabolites with a log_2_ fold change >1 and adjusted p < 0.05 are labeled and colored (with a log_2_ fold change > 0.5 only colored) based on pathways or molecular families as shown in the key. **C.** Abundances of metabolites in the tryptophan pathway were compared between VD and CS mice using Student’s t-test. For each metabolite, log_2_ fold change and p-value were calculated and mapped as shown. Red and blue circles indicate increases and decreases in VD mice in comparison to CS mice respectively. The size of each circle reflects statistical significance. **D-E**. z-scale values of histamine and acetylcholine (D) and arginine cycle metabolites (E). Each dot represents a mouse and is coded as shown in the key. Statistical signifcance was assessed by one-way ANOVA with Tukey’s multiple comparison test and only statistically signficant comparisons are shown (not shown, p > 0.05; * p < 0.05; ** p < 0.01; ***p < 0.001; ****p < 0.0001).

Tryptophan metabolism shifted in VD mice towards consumption of tryptophan in a non-specific manner by all three major routes of enzymatic tryptophan conversion, namely the serotonin, kynurenine and indole pathways (Figure 4C). In comparison, tryptophan and indole were elevated in CS and CS+VS suggesting decreased metabolic activity (Figure 4B-C, Figure S5A). Interestingly, serotonin and the kynurenine-metabolite picolinic acid trended downward in CS+VS mice compared to CS mice (Figure S7). Histamine was increased in colonized mice compared to germ-free controls, suggesting a general dependance on microbes (Figure 4D, Figure S5A). Further, histamine was significantly elevated in VD mice in comparison to CS+VS mice, with CS mice displaying the highest levels across the groups albeit significant variation within this group (Figure 4D). In contrast to the histamine pattern, acetylcholine was significantly elevated only in VD mice (Figure 4D).

Amine metabolism was activated in VD mice fecal metabolome with concomitant decrease in arginine and increases in arginine cycle metabolites citrulline, ornithine, ADMA and SDMA, and downstream polyamines, putrescine and spermidine (Figure 4B, 4E, Figure S6). Interestingly, ornithine and citrulline levels were highest in the metabolome of CS+VS mice across the groups (Figure 4B, 4E). However, CS+VS mice did not show differences in ADMA and SDMA and failed to activate polyamine production downstream of the arginine cycle with decreased levels in both putrescine and n-acetylputrescine (Figure 4E, Figure S7). These findings suggest that in comparison to CS mice, CS+VS mice are able to partially activate the amine metabolism cycle.

Flavin and nicotinamide cofactor metabolism displayed significant activation in VD mice with a ubiquitous increase in all measured metabolites in both pathways compared to CS and CS+VS mice (Figure 4B, Figure 5A, Figure S6). The only effect of VS as observed from CS+VS mice was elevated levels of nicotinamide in comparison to CS mice (Figure S7). In terms of central energy metabolism, VD mice displayed increased glycolytic intermediates and TCA cycle metabolites whereas CS and CS+VS mice shared the common feature of enrichment in certain short-chain fatty acyl-carnitines (Figure S5A, S6, S7). Thus, our findings from fecal metabolomic analyses revealed that the VD microbiome associated with strong activation of key homeostatic metabolic pathways, with CS+VS microbiome colonized mice displaying distinct differences in amine metabolism.

**Figure 5.**
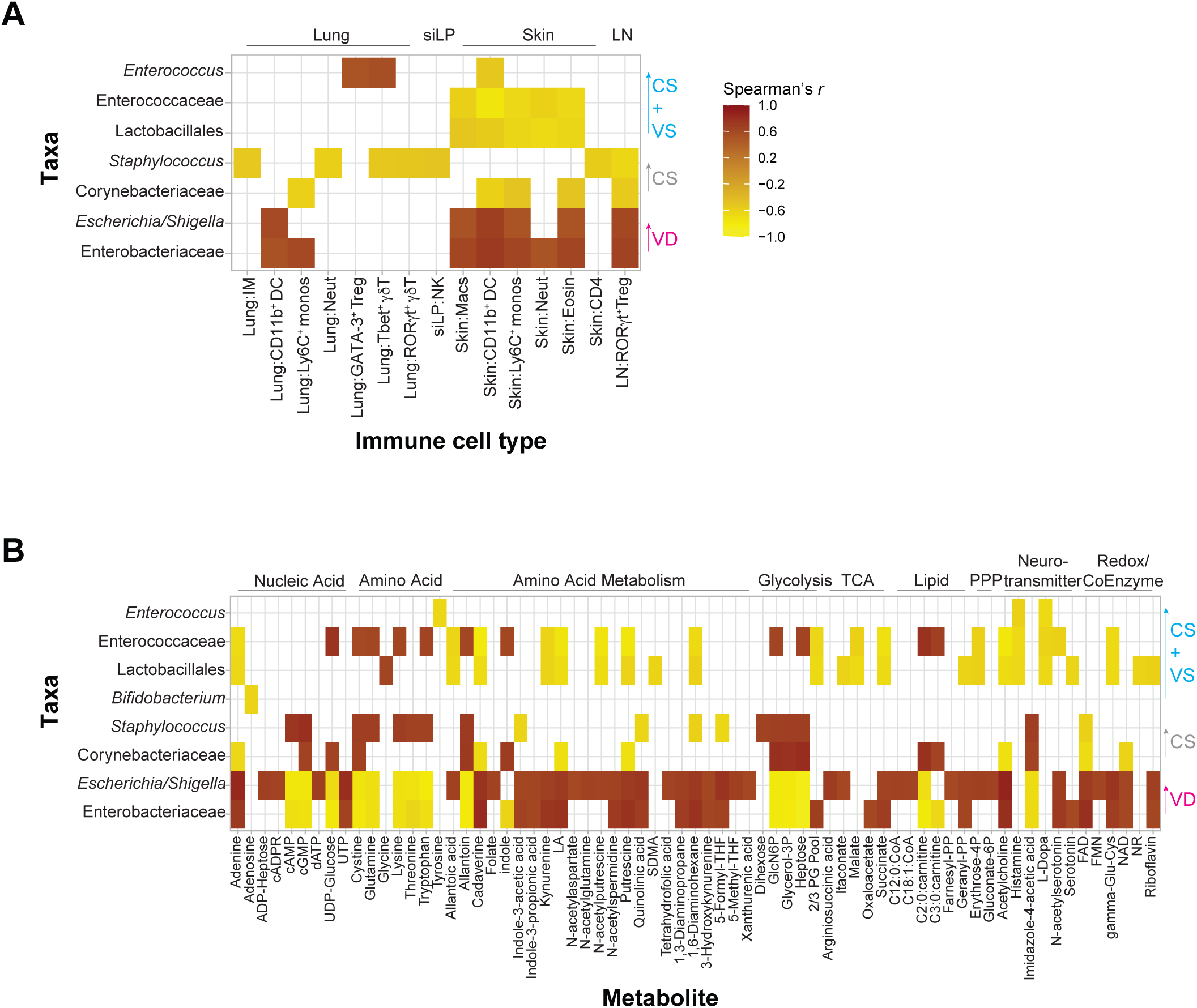
Correlation analysis between microbial taxa, metabolites and immune cell types identified in colonized germ-free mice. **A-B** Spearman’s correlation coefficient between measurements of significantly altered fecal microbial taxa and immune cell types (A) and fecal microbial taxa and fecal metabolites (B) were calculated. Correlations were filtered for correlation coefficient r > |0.5| and adjusted p < 0.05 (A) or r > |0.6| and adjusted p < 0.01 (B) and displayed as a heatmap. Correlations shown are based on measurements from all subjects across the three groups of colonized mice. Taxa identified at order or family level indicate unclassified genera under those taxa, as represented in Figure 1E.

### Correlative analysis identified microbial taxa associated with immune and metabolic changes

Finally, we performed correlative analyses to identify taxa that potentially drive the observed immune and metabolic profiles. We found that taxa that were enriched in the VD mice, *Escherichia/Shigella* and an Enterobacteriaceae genus, positively correlated with myeloid cells increased in the lung and skin, with RORγt^+^ Tregs in the LN being the only significant positive correlation to a lymphoid cell type (Figure 5A). These same taxa showed significant correlations with key metabolic patterns discussed above (Figure 5B). These include flavin and nicotinamide cofactors, acetylcholine, succinate, polyamines and activation of tryptophan metabolism toward indole, serotonin, and kynurenine products (Figure 5B). These taxa also negatively correlate with several amino acids and nucleic acids. Further, the metabolites elevated in VD mice also positively correlate with the immune cell types noted above (Figure S8).

In contrast, taxa that define the CS microbiome namely, a Corynebacteriaceae genus and *Staphylococcus*, exhibit significant negative correlations to myeloid cells in the lung and skin, lung γδ T cell subsets, skin CD4^+^ T cells and LN RORγt^+^ Tregs and show positive correlations with glycolysis-adjacent metabolites glycerol-3P and GlcN6P and the diamine oxidase histamine metabolite imidazole-4-acetic acid (Figure 5A-B). The correlation patterns of taxa elevated in CS+VS mice follow a similar trend to that observed with CS mice with the notable exception being a positive correlation between *Enterococcus* and, GATA-3+ Tregs and Tbet^+^ γδ T cells in the lung (Figure 5A-B). Overall, these analyses demonstrate that the specific taxa that colonize the mice drive distinct immune and metabolic environments, and these are significantly different between VD and CS groups.

## DISCUSSION

CS is a life-saving surgery that is often medically necessary. Currently, 21% of live births worldwide and 32% in the United States occur via CS^41,42^. Given the high rate of CS births and increasing evidence of CS associated disease risks that is at least in part attributed to a different microbiome acquired at birth, it is essential to mechanistically better understand, how the altered microbiome affects host health. Such investigations will aid in the development of intervention strategies aimed at mitigating CS associated health impacts.

Here, utilizing a human-to-murine fecal transplant model we demonstrated that germ-free mice colonized with CS acquired microbiome in comparison to those associated with VD microbiota display decreased myeloid and regulatory T cells, a Th2 skew and decreased microbial metabolic activity. Additionally, we observe this distinct modulation of the immune environment not only in the gut, but also in extra-intestinal barrier tissues, namely the lung and skin, indicating a systemic influence of the intestinal microbiome and metabolome. Importantly, the microbiome from VS neonates is able to partially restore this phenotype as evidenced by increased regulatory T cells, serum IgA and a slightly decreased Th2 bias, but does not greatly recover metabolic activity.

The immune system in germ-free mice is underdeveloped but can be rescued with colonization of a defined consortia of microbes early in life ^3,5,43,44^. In a similar context, a newborn infant shows immune immaturity with a Th2 bias including increased IgE and diminished pro-inflammatory responses, which are important to prevent overt inflammation during microbial colonization and subsequent immune education^16,31,45^. By utilizing pioneering colonizers that are reflective of microbes that the infant is seeded with during birth, to the best of our knowledge, we have for the first time, demonstrated that different clinical microbiomes acquired as a consequence of delivery mode drive differential modulation of the immune and metabolic environments. This, in turn, can potentially set the tone of the trajectory of immune maturation. Previous studies have reported a prolonged Th2 bias and delayed immune maturity in CS delivered infants compared to VD infants^16,31^. Our findings emphasize that the rewiring to a non-allergic environment and immune maturation starts from birth and is directly influenced by the pioneering microbes acquired as a consequence of delivery mode.

The most significant microbial signature in our VD mice is the enrichment of *Escherichia* and another genus of family Enterobacteriaceae. Members of this taxa are facultative anaerobes and are often the first colonizers of the infant gut that can condition the intestinal environment for consequent colonization with obligate anaerobes such as *Bifidobacterium* and *Bacteroides*^46,47^. Further, they express LPS and flagellin that activate the innate immune system via toll-like receptors and shape mucosal responses and barrier integrity^48–51^. In line with this, we observed an increased presence of myeloid cells including monocytes and dendritic cells in VD mice. Continued microbial colonization can lead to dendritic cell subset differentiation that is critical for helper T cell differentiation and induction of Tregs^52^.

The introduction of the pioneering microbiome also begins the maturation of metabolic homeostasis in the neonate. Here we show that this metabolic maturation can be distinct based on specific microbiota community. The VD microbiome is associated with modulation of metabolic pathways known to be involved in immune-metabolic homeostasis in the gut including tryptophan metabolites from the indole, serotonin and kynurenine pathways, increased coenzyme and acetylcholine levels and shifts in energy metabolism^7,53–55^. Histamine and the arginine cycle metabolites displayed differential modulation across the VD, CS and CS+VS microbiota communities. Histamine’s role in the gastrointestinal tract is multifaceted including regulation of pH, smooth muscle function and immunoregulation^55^. While histamine is elevated following microbial colonization, the level of increase and the specific degradation pathways that are active are species dependent. Similarly, activation of the arginine cycle, as represented by levels of citrulline and ornithine, was a shared feature of colonization and suggests increased activity of protective Endothelial Nitric Oxide Synthase (eNOS)^56^. However, production of secondary arginine metabolites was specific to VD mice, which displayed higher fecal levels of the eNOS-regulating dimethylarginines and the barrier-regulating polyamines^57,58^. Metabolites that characterized the VD metabolic pattern correlated with specific taxa in the VD microbiome, specifically *Escherichia* and an other Enterobacteriaceae genus. *Escherichia coli* has been previously shown to synthesize indole acids including indole-3-lactic acid which activates AhR pathways, promotes Treg differentiation, and anti-inflammatory responses in immature epithelial cells^59–62^. These same taxa were correlated with polyamines and *E. coli* and other Enterobacteriaceae synthesize putrescine and spermidine, that is absorbed by colonocytes to improve barrier function and regulate macrophage activity^53^. The fecal metabolome of CS mice lacks these metabolites that perform key functions in metabolic homeostasis and is instead enriched in unmetabolized amino acid substrates suggesting that the CS microbiota potentially lack this metabolic capacity.

An additional objective of this study was to examine the potential restorative immune and metabolic benefits of vaginal seeding. In mice colonized with microbiome from CS+VS neonates, we observed increased Tregs, Th1/Th2 ratio and serum IgA. The increased Treg population may be explained by the enrichment of a Lactobacillales genus and *Bifidobacterium* in the CS+VS microbiome. Interestingly, a recent study demonstrated that CS infants that harbor fewer maternal-acquired microbes are colonized with higher level of breast milk acquired microbes, such as *Lactobacillus* and *Bifidobacterium*, in comparison to VD infants^13^. Metabolically, CS+VS mice closely resembled CS mice with isolated differences in arginine cycle metabolites, potentially indicative of differences in barrier function and in the degree of activation of the eNOS system discussed above^63^. The myeloid compartments in CS+VS and CS mice were similar, overall, indicating only a partial benefit of VS. Perhaps this finding is not too surprising, given that microbial taxa, *Escherichia* and an other Enterobacteriaceae genus, that were key drivers of the phenotype, observed in VD mice were absent in CS+VS mice. However, in a previous study we were able to identify *Escherichia* in mice colonized with stool from CS+VS infants suggesting variability among study subjects and efficacy of the VS procedure^40^. Nevertheless, these findings raise the possibility of defining and optimizing a consortium of microbes that may be used for VS of CS delivered infants that sets a baseline immune tone similar to VD babies. It should be noted that *Escherichia*, identified here as a primary driver of a beneficial phenotype, has been linked to adverse outcomes in pre-term infants with several strains being pathogenic, and thus this promising avenue of research warrants further extensive critical investigation^64^.

There are limitations in this study and areas that would benefit from additional investigation. First, the study focused on bacterial microbes, however, the importance of other members of microbiota, such as fungi, in setting the baseline immune and metabolic tone needs to be examined. Further, only the significance of the intestinal microbiome in these animals was assessed; the resident microbes at other barrier sites could also be relevant and need to be explored. Additionally, insufficient material was available to perform sequencing of the infant stool used for inoculating mice, thereby precluding a comparative analysis of the microbial composition between the original human stool samples and the corresponding murine fecal samples. Second, while the pioneering colonizers may set the tone of immune and metabolic development, some of these taxa particularly Enterobacteriaceae are only transiently abundant. The impact of the initial colonizers on defining the subsequent colonization and modulation of the mucosal environment, and how this determines health outcomes remain to be investigated. Further, in this study mice were colonized at weaning; colonization at birth may have resulted in more pronounced differences. Therefore, future experiments should investigate the effects of colonization initiated at birth. Third, unlike laboratory mice, humans exhibit high interindividual variability which can be particularly true in case of interventional studies as we have observed in our cohort of VS trial subjects in this current study and previous study^40^. Human variability limits the feasibility and broader applicability of human-to-murine fecal transplant models for mechanistic investigations. A priori knowledge of the microbial composition and any health outcomes of subjects during sample selection may provide more meaningful mechanistic insights and biological significance.

## METHODS

### EXPERIMENTAL MODEL AND SUBJECT DETAILS

#### Human Subjects

Stool samples from VD infants were obtained from an Institutional Review Board (IRB) approved clinical study (WCG protocol #20120204) in which pregnant women were enrolled after informed consent. Stool samples from their infants including from day 2 or 3 of life were collected and stored at -80°C. Subjects in the CS and CS+VS cohorts were enrolled in an IRB approved (WCG protocol #1300043) randomized controlled clinical trial where infants born via CS were randomized to receive a “wipe-down” with saline gauze (CS group) or gauze with mother’s own vaginal fluid (CS+VS group) as previously described^38^. Briefly, on the day of the scheduled CS prior to any preoperative antibiotics, a saline moistened gauze was inserted into the mother’s vagina, incubated for approximately 1 hour and stored in a sterile container (VS gauze). Immediately following CS delivery, prior to skin-to-skin contact with the mother, the infant was wiped down with either the VS or only saline moistened gauze, depending on the randomization. Stool samples from day 2 or 3 were frozen in -80°C. Detailed maternal and infant clinical characteristics and demographics were collected for both clinical studies. Samples chosen for murine studies were based on availability.

#### Mice

Germ-free C57Bl/6NTac mice were bred and maintained at the NIAID Gnotobiotic animal facility and all procedures were performed in accordance with the Animal Study Proposal LHIM 2E approved by the NIAID Animal Care and Use Committees. Mice were routinely screened via microbiological culture and *16S rRNA* PCR to ensure their germ-free status prior to inoculation. Mice were inoculated with one human infant stool at the time of weaning and weaned into separate cages by sex.

## METHOD DETAILS

### Colonization of germ-free mice with human stool

Infant stool collected on day 2 or 3 of life and stored at -80°C was used to prepare the inoculum to colonize germ-free mice. The stool sample was resuspended in sterile pre-reduced PBS at a concentration of ∼620 mg/ml in an anerobic chamber and stored in a Hungate tube until inoculation. At the time of weaning each mouse was orally gavaged with 100 μl of the stool slurry and housed in cages by sex in biocontainment racks. Experiments were performed independently for each human subject. Stool from each subject was inoculated into a combination of 4-9 male and female mice. Downstream data were not analyzed by sex as the study was not sufficiently powered for such analyses.

### *16S rRNA* variable region amplicon sequencing and microbiome analysis

Fecal material was homogenized in Lysis Matrix E tubes (MP Biomedicals) with 650 μl MBL lysis buffer (MagAttract PowerMicrobiome DNA/RNA EP kit, Qiagen) using a Precellys 24 Tissue Homogenizer (Bertin Technologies). Homogenized samples were centrifuged, and DNA was purified from the supernatant using the MagAttract PowerMicrobiome DNA/RNA EP Kit on an automated liquid handling system (Eppendorf) as per the manufacturer’s instructions. For *16S rRNA* sequencing, the V4 region was amplified using 515F and 806R primers as previously described^65^. Briefly, PCR amplification was performed using 0.2 μM of F/R primers, 1X Phusion High-Fidelity DNA Polymerase (New England Biolabs) and 4 ng of fecal DNA with the following cycling conditions: initial template denaturation at 98°C for 60 seconds; 25 cycles of denaturation at 98°C for 10 seconds, primer annealing at 55°C for 30 seconds and template extension at 72°C for 60 seconds and a final template extension at 72°C for 5 min. PCR products were purified using AMPure XP beads (Beckman-Coulter) at a ratio of 1:1 and the libraries were quantified using the KAPA qPCR Library Quantification Kit (Kapa Biosystems) and pooled at an equimolar concentration for sequencing. Paired-end sequencing (250×250) was performed using MiSeq Reagent Kit v3 (600 cycles) on the Illumina MiSeq platform, with a 15% PhiX control library spike-in.

The sequenced demultiplexed paired-end reads were processed and analyzed using QIIME2/DADA2 pipeline version 2-2024.2^66,67^. Raw reads were first trimmed and filtered based on sequencing quality. After denoising and chimera removal, 18,188,974 reads were obtained from 216 samples consisting of small intestine and colon fecal contents and fecal pellets. Following analysis of alpha rarefaction curves, samples were rarefied at a sampling length of 2,000 reads for diversity analyses given the low microbiota diversity typically observed in transitional stool. 23 out of 216 samples, from 8 mice, had less than 2000 sequencing reads and were removed from all further analyses. Alpha-diversity metrics, observed ASVs and Shannon, and beta diversity Bray-Curtis distance matrix were calculated using the phyloseq package in R^68^. Kruskal-Wallis test and Adonis were used to assess statistical significance for alpha and beta diversity measurements respectively. Taxonomic classification was performed using Silva database release 138^69^. ASVs that were not classified with a specific genus name were identified as an unclassified genus under the highest taxonomic classification reported for differential abundance analysis. ANCOMBC in QIIME2 was used to identify differentially abundant taxa in pairwise comparisons of experimental groups for each fecal sampling site and taxa with q value < 0.05 following multiple testing were considered statistically significant.

### Tissue processing

Mice were euthanized using CO_2_ and blood was collected by cardiac exsanguination. Mesenteric lymph node, small intestine, colon, lung and one ear pinnae (for skin) were harvested. Small intestine and colon were flushed with 5 ml of pre-reduced PBS to collect the fecal contents that were stored at -80°C. For siLP preparation, Peyer’s patches and adipose tissue were removed, the tissue opened longitudinally and washed in ice cold PBS to remove any additional feces. The tissue was then cut into 1-2 cm pieces and incubated in complete media (RMPI 1640 supplemented with 20 mM HEPES, 2 mM L-glutamine, 1 mM sodium pyruvate, 1 mM nonessential amino acids, 50 mM β-mercaptoethanol, 100U/ml penicillin and 100 mg/ml streptomycin) + 3% FBS + 5 mM EDTA + 0.145 mg/ml DL-dithiothreitol for 20 min at 37°C and 5% CO_2_ with shaking. Tissues were then vigorously shaken in a 50 ml tube containing 10 ml complete media + 2mM EDTA and strained thrice, to remove epithelial cells, and finely chopped and digested with 10 ml of digestion media (complete media + 0.1 mg/ml Liberase TL+ 0.05% DNase I, both from Sigma-Aldrich) for 25 min at 37°C and 5% CO_2_ with shaking. The reaction was stopped with 10 ml of complete media + 3% FBS. The digested tissues were passed through a 100 μm strainer and then 40 μm strainer with centrifugation after each straining step to isolate siLP cells.

Lung lobes were chopped into small pieces and digested in RMPI 1640 + 0.33 mg/ml Liberase TL + 0.1 mg/ml DNase I at 37°C for 40 min with shaking. The digestion reaction was stopped with FBS and tissues passed through a 70 μm strainer. Red blood cells were lysed using ACK lysing buffer and lungs cells were pelleted via centrifugation. To isolate skin cells, excised ear pinnae were separated, and the dermal side was placed facing down on the digestion media containing complete media + 0.25 mg/ml Liberase TL+ 0.5 mg/ml DNase I and incubated for 90 min at 37°C. The digested ear tissues were then homogenized using a Medimachine tissue homogenizer and passed through a 50 μm filter and centrifuged to generate single cell suspensions. Mesenteric lymph nodes were passed through a 70 μm strainer and centrifuged. All centrifugation steps were performed at 1500 rpm for 5 min at 4°C, and all isolated single cells were resuspended in RPMI + 10% FBS prior to flow cytometry analysis.

### Spectral flow cytometry

For analysis of intracellular lymphoid cell cytokine production potential, cells were stimulated with Cell Stimulation cocktail 500X (ThermoFisher) prepared in complete media + 10% FBS for 2.5 hours at 37°C. To assess lymphoid and myeloid cells, cells were incubated with Zombie NIR Fixability Dye and TruStain FcX (both from Biolegend) for 15 min in the dark, following which cells were incubated with cocktails of fluorescently conjugated antibodies for surface markers diluted in PBS + 1% FBS + 10% Brillant Stain Buffer for 25 min in the dark at 4°C. Cells were then fixed overnight with eBioscience Fixation/Permeabilization Solution Kit (Invitrogen) at 4°C. For intracellular antigens and cytokines, a cocktail of fluorescently labeled antibodies was prepared in eBioscience intracellular staining buffer (Invitrogen) and cells were stained for 40 min at room temperature. Cells were then washed twice, and data for all samples were collected on a spectral cytometer (Cytek Aurora). Spectral unmixing was performed using single-stained controls of cells from corresponding tissues or UltraComp Beads (Invitrogen). Data were analyzed using FlowJo version 10.

### Detection of serum antibodies

Serum IgA and IgE concentrations were determined using Mouse IgA (ThermoFisher) and Mouse IgE (BioLegend) ELISA Kits respectively, following manufacturer’s protocol.

### Fecal liquid chromatography tandem mass spectrometry (LC-MS/MS) based metabolomics

To each frozen fecal pellet, 400 ul methanol was added followed by 400 ul water and 400 ul chloroform. Samples were agitated for 30 min at 4°C then centrifuged at 16,000*g* for 20 min. 200 ul of the aqueous layer was collected and diluted 5X in 50% methanol, combined 4:1 with LC-MS/MS buffer A below prior to injection. Aqueous metabolites were separated using a Hydrophilic Interaction Liquid Chromatography method on a LD40 XR UHPLC (Shimadzu Co.) system and detected using a 6500+ QTrap mass spectrometer (AB Sciex Pte. Ltd.). A Water XBridge Amide column (3.5 μm, 3 mm X 100 mm) and metabolites were eluted using a binary gradient from A: 5% 5mM aqueous ammonium acetate pH 7.5 95% acetonitrile to B: 95 % 5 mM aqueous ammonium acetate pH 7.5 and 5% acetonitrile over 6.75 minutes. Multiple reaction monitoring (MRM) ion pairs were taken from a variety of previously published methods and fidelity assessed using in-house murine organ standards^70,71^. Quality control samples for assessment of instrument stability were injected every 10 injections. All signals were integrated using SciexOS 3.1 (AB Sciex Pte. Ltd.). Signals with greater than 50% missing values were discarded and remaining missing values were replaced with the lowest registered signal value. Where appropriate, signals with a QC coefficient of variance greater than 30 % were discarded. Metabolites with multiple MRMs were quantified with the higher signal to noise MRM. Filtered datasets were total sum normalized after initial filtering.

### Quantitative and statistical analysis

Statistical analyses for microbiome data were performed as described above. Statistical significance for flow cytometry and serum antibody measurements was assessed using one-way ANOVA. Principal components plots were generated using the FactoMineR package in R. To determine significantly altered metabolites across all groups, ANOVA with multiple-testing correction was performed and visualized as a heatmap using pheatmap function in R. To identify, differentially abundant metabolites in pairwise comparisons between groups, Student’s t-test with multiple-testing correction was used. Average log_2_ fold change of the metabolites with adjusted p-values were plotted using EnhancedVolcano package in R. Metabolic pathway maps were generated in Cytoscape. Pairwise spearman’s correlation analyses with multiple-testing correction were performed to identify significant correlations between taxa, metabolites and immune readouts and plotted as a heatmap using ggplot in R. Benjamini-Hochberg method was used for all multiple testing corrections. Statistically significant cut-offs used in each analysis are indicated in the figure legends.

## DATA AVAILABILITY

Microbiome sequencing data generated in this study has been submitted to NCBI’s Short Read Archive Database under submission PRJNA1330279. The metabolomics data has been submitted to Figshare under https://doi.org/10.6084/m9.figshare.30209614.v1

## ACKNOWLEDGEMENTS

We thank the NIAID Gnotobiotic Animal Facility, Ejae Lewis and Dr. Nicolas Bouladoux for support in performing the germ-free experiments. We thank Joshua Gold for his help with metabolomics sample extraction and Nehamiah Strawberry and Avery D. Walters for their help with generation of metabolomics figures. We also acknowledge the NIH HPC Biowulf cluster for providing computation resources. We thank Dr. Dragana Jankovic for critical reading of the manuscript. The authors thank Kristy and Roger Crombie for their generous philanthropic donation toward this project in loving memory of their daughter Anna Charlotte. This research was supported by the Intramural Research Program of the National Institutes of Health (NIH). The contributions of the NIH author(s) were made as part of their official duties as NIH federal employees, are in compliance with agency policy requirements, and are considered Works of the United States Government. However, the findings and conclusions presented in this paper are those of the author(s) and do not necessarily reflect the views of the NIH or the U.S. Department of Health and Human Services.

## AUTHOR CONTRIBUTIONS

SN, YB and SKH conceived the study and established the mouse model. SN, HNR, MB, PJP, PL and KB performed the murine experiments. ASB and SM performed the microbiome sequencing. NTB, AC, GC, ISL and BS performed the metabolomics. QC, AP, NTM, MGD, GLM, SL and SKH provided clinical samples. SN analyzed the data and generated the figures. SN and SKH wrote the manuscript and all authors reviewed and edited the manuscript.

## SUPPLEMENTAL FIGURE LEGENDS

**Figure S1. Relative abundance of bacterial taxa in mice colonized with stool from neonates born via different delivery modes**

**A.** Relative abundance of bacterial families in the intestinal microbiota of colonized mice. Data are grouped by infant stool donor across the three study groups, with each stacked bar representing an individual mouse. **B.** Comparison of relative abundances of taxa identified as significantly different between groups in Figure 1E.

**Figure S2. Gating strategy to identify immune cell types by spectral flow cytometry**

**A-B.** Flow cytometry plots depict the gating strategy of myeloid (A) and lymphocyte (B) subsets in the siLP, lung, skin and mesenteric LN (lymphocyte only). Gating for tissue specific myeloid subsets are also indicated.

**Figure S3. Comparison of major lymphoid cell types in colonized mice**

**A-B.** Quantification of innate lymphocyte subsets in siLP (A) and lung (B). **C-D.** Frequency CD4^+^ T cell subsets in siLP (C) and lung (D). **E-F.** Frequency of γδ T cell subsets in siLP (E) and lung (F). Each dot represents an individual mouse and is coded as shown in the key. Statistical signifcance was assessed by one-way ANOVA with Tukey’s multiple comparison test and only statistically signficant comparisons are shown (not shown, p > 0.05; * p < 0.05; ** p < 0.01; ***p < 0.001; ****p < 0.0001)

**Figure S4. Comparison of lymphoid cell types in mesenteric lymph nodes of mice colonized with neonatal stool**

**A.** Cell numbers of major lymphocyte cell types. **B-C.** Number (B) and frequency (C) of CD4^+^ T cell subsets**. D.** Ratio of Th1/Th2 and Th1/Th17 cells. **E-F.** Frequency of IFNγ^+^, IL-17A^+^ and IL-22^+^ CD4^+^ T cells (E) and IFNγ^+^ CD8^+^ T cells (F) following ex-vivo stimulation **G.** Frequency of γδ T cell subsets. **H.** Frequency of IFNγ^+^ and IL-17A^+^ γδ T cells following ex-vivo stimulation. **I.** Ratio of Tbet^+^/RORγt^+^ γδ T cells. Each dot represents an individual mouse and is coded as shown in the key. Statistical signifcance was assessed by one-way ANOVA with Tukey’s multiple comparison test and only statistically signficant comparisons are shown (not shown, p > 0.05; * p < 0.05; ** p < 0.01; ***p < 0.001; ****p < 0.0001)

**Figure S5. Differentially abundant fecal metabolites across colonized and control mice**

**A.** Significantly altered metabolites among the three study groups and vehicle control group were identified using one-way ANOVA and filtered for adjusted p < 0.0001. Normalized metabolite signals are displayed as a heatmap with each row representing a metabolite grouped by pathways or families. Each column represents data from one mouse color-coded by group with color gradients indicating different human donor subjects used to inoculate the mice. **B.** Volcano plots show fecal metabolites that were differentially abundant in pairwise comparisons between each study group and vehicle treated control group as indicated. Statistical significance was assessed using Student’s t-test and metabolites with a log_2_ fold change >1 and adjusted p < 0.05 are labeled and colored (with log2 fold change > 0.5 only colored) based on pathways or molecular families as shown in the key.

**Figure S6. Differential abundance of fecal metabolites between VD and CS mice mapped to metabolic pathways**

Fecal metabolite abundances between VD and CS mice were compared by Student’s t-test. Log_2_ fold change and corresponding p-values were calculated and mapped to metabolites within pathways, as shown. Red and blue circles indicate increases and decreases in VD mice in comparison to CS mice respectively. The size of each circle reflects statistical significance.

**Figure S7. Differential abundance of fecal metabolites between CS+VS and CS mice mapped to metabolic pathways**

Fecal metabolite abundances between CS+VS and CS mice were compared by Student’s t-test. Log_2_ fold change and corresponding p-values were calculated and mapped to metabolites within pathways, as shown. Red and blue circles indicate increases and decreases in CS+VS mice in comparison to CS mice respectively. The size of each circle reflects statistical significance.

**Figure S8. Correlation between metabolites and immune cell types in colonized mice**

Spearman’s correlation coefficient between measurements of immune cell types and significantly altered fecal metabolites were calculated and correlations with r > |0.5| and adjusted p < 0.05 are displayed as a heatmap. Correlations shown are based on measurements from all subjects across the three groups of colonized mice.

**Supplemental Table 1:** Infant and maternal clinical characteristics of human subjects.

|  | **Vaginally**<br>**delivered**<br>**(VD)**<br>**(n=4)** | **Cesarean**<br>**section**<br>**delivered**<br>**(CS)**<br>**(n=3)** | **Cesarean**<br>**delivered and**<br>**vaginally**<br>**seeded (CS+VS)**<br>**(n=3)** |
| --- | --- | --- | --- |
| **Infant Characteristics** |  |  |  |
| Gestational age in weeks (mean) | 38.95 | 39 | 38.67 |
| Breast milk prior to stool sample, n (%) | 4 (100%) | 3 (100%) | 3 (100%) |
| Formula prior to stool sample, n (%) | 2 (50%) | 0 (0%) | 1 (33%) |
| Antibiotics prior to stool sample, n (%) | 0 (0%) | 0 (0%) | 0 (0%) |
| Sex female, n (%) | 2 (50%) | 2 (67%) | 0 (0%) |
| **Maternal Characteristics, n (%)** |  |  |  |
| Pre-pregnancy body mass index |  |  |  |
| < 25 | 2 (50%) | 3 (100%) | 1 (33%) |
| 25-30 | 2 (50%) | 0 (0%) | 1 (33%) |
| >30 | 0 (0%) | 0 (0%) | 1 (33%) |
| Prenatal Antibiotics | 0 (0%) | 0 (0%) | 0 (0%) |
| Peripartum Antibiotics | 0 (0%) | 3 (100%) | 3 (100%) |
| Race |  |  |  |
| White | 3 (75%) | 3 (100%) | 1 (33%) |
| Black or African American | 0 (0%) | 0 (0%) | 1 (33%) |
| More than one race/Other/Decline to answer | 1 (25%) | 0 (0%) | 1 (33%) |
| Ethnicity |  |  |  |
| Hispanic or Latino | 1(25%) | 0 (0%) | 1 (33%) |
| Not Hispanic or Latino/Other | 3 (75%) | 3 (100%) | 2 (67%) |

## REFERENCES

1. Donald, K. & Finlay, B.B. Early-life interactions between the microbiota and immune system: impact on immune system development and atopic disease. Nat Rev Immunol 23, 735–748 (2023).

2. Jian, C., et al. Early-life gut microbiota and its connection to metabolic health in children: Perspective on ecological drivers and need for quantitative approach. EBioMedicine 69, 103475 (2021).

3. Gensollen, T., Iyer, S.S., Kasper, D.L. & Blumberg, R.S. How colonization by microbiota in early life shapes the immune system. Science 352, 539–544 (2016).

4. Brodin, P. Immune-microbe interactions early in life: A determinant of health and disease long term. Science 376, 945–950 (2022).

5. Cahenzli, J., Koller, Y., Wyss, M., Geuking, M.B. & McCoy, K.D. Intestinal microbial diversity during early-life colonization shapes long-term IgE levels. Cell Host Microbe 14, 559–570 (2013).

6. Round, J.L. & Mazmanian, S.K. Inducible Foxp3+ regulatory T-cell development by a commensal bacterium of the intestinal microbiota. Proc Natl Acad Sci U S A 107, 12204–12209 (2010).

7. Constantinides, M.G., et al. MAIT cells are imprinted by the microbiota in early life and promote tissue repair. Science 366(2019).

8. Cox, L.M., et al. Altering the intestinal microbiota during a critical developmental window has lasting metabolic consequences. Cell 158, 705–721 (2014).

9. Bokulich, N.A., et al. Antibiotics, birth mode, and diet shape microbiome maturation during early life. Sci Transl Med 8, 343ra382 (2016).

10. Tamburini, S., Shen, N., Wu, H.C. & Clemente, J.C. The microbiome in early life: implications for health outcomes. Nat Med 22, 713–722 (2016).

11. Milani, C., et al. The First Microbial Colonizers of the Human Gut: Composition, Activities, and Health Implications of the Infant Gut Microbiota. Microbiol Mol Biol Rev 81(2017).

12. Sprockett, D., Fukami, T. & Relman, D.A. Role of priority eaects in the early-life assembly of the gut microbiota. Nat Rev Gastroenterol Hepatol 15, 197–205 (2018).

13. Bogaert, D., et al. Mother-to-infant microbiota transmission and infant microbiota development across multiple body sites. Cell Host Microbe 31, 447–460 e446 (2023).

14. Reyman, M., et al. Impact of delivery mode-associated gut microbiota dynamics on health in the first year of life. Nat Commun 10, 4997 (2019).

15. Chu, D.M., et al. Maturation of the infant microbiome community structure and function across multiple body sites and in relation to mode of delivery. Nat Med 23, 314–326 (2017).

16. Lai, C., et al. Eaect of diaerent delivery modes on intestinal microbiota and immune function of neonates. Sci Rep 14, 17452 (2024).

17. Li, N., et al. Distinct gut microbiota and metabolite profiles induced by delivery mode in healthy Chinese infants. J Proteomics 232, 104071 (2021).

18. Wampach, L., et al. Birth mode is associated with earliest strain-conferred gut microbiome functions and immunostimulatory potential. Nat Commun 9, 5091 (2018).

19. Sevelsted, A., Stokholm, J., Bonnelykke, K. & Bisgaard, H. Cesarean section and chronic immune disorders. Pediatrics 135, e92–98 (2015).

20. Huh, S.Y., et al. Delivery by caesarean section and risk of obesity in preschool age children: a prospective cohort study. Arch Dis Child 97, 610–616 (2012).

21. Zachariassen, L.F., et al. Cesarean Section Induces Microbiota-Regulated Immune Disturbances in C57BL/6 Mice. J Immunol 202, 142–150 (2019).

22. Vu, K., et al. From Birth to Overweight and Atopic Disease: Multiple and Common Pathways of the Infant Gut Microbiome. Gastroenterology 160, 128–144 e110 (2021).

23. Slabuszewska-Jozwiak, A., Szymanski, J.K., Ciebiera, M., Sarecka-Hujar, B. & Jakiel, G. Pediatrics Consequences of Caesarean Section-A Systematic Review and Meta-Analysis. Int J Environ Res Public Health 17(2020).

24. Darmasseelane, K., Hyde, M.J., Santhakumaran, S., Gale, C. & Modi, N. Mode of delivery and oaspring body mass index, overweight and obesity in adult life: a systematic review and meta-analysis. PLoS One 9, e87896 (2014).

25. Kuhle, S., Tong, O.S. & Woolcott, C.G. Association between caesarean section and childhood obesity: a systematic review and meta-analysis. Obes Rev 16, 295–303 (2015).

26. Li, H.T., Zhou, Y.B. & Liu, J.M. The impact of cesarean section on oaspring overweight and obesity: a systematic review and meta-analysis. Int J Obes (Lond) 37, 893–899 (2013).

27. Dominguez-Bello, M.G., et al. Delivery mode shapes the acquisition and structure of the initial microbiota across multiple body habitats in newborns. Proc Natl Acad Sci U S A 107, 11971–11975 (2010).

28. Shao, Y., et al. Stunted microbiota and opportunistic pathogen colonization in caesarean-section birth. Nature 574, 117–121 (2019).

29. Wong, W.S.W., et al. Prenatal and Peripartum Exposure to Antibiotics and Cesarean Section Delivery Are Associated with Diaerences in Diversity and Composition of the Infant Meconium Microbiome. Microorganisms 8(2020).

30. Zhang, C., et al. The Eaects of Delivery Mode on the Gut Microbiota and Health: State of Art. Front Microbiol 12, 724449 (2021).

31. Jakobsson, H.E., et al. Decreased gut microbiota diversity, delayed Bacteroidetes colonisation and reduced Th1 responses in infants delivered by caesarean section. Gut 63, 559–566 (2014).

32. Backhed, F., et al. Dynamics and Stabilization of the Human Gut Microbiome during the First Year of Life. Cell Host Microbe 17, 852 (2015).

33. Hourigan, S.K., Dominguez-Bello, M.G. & Mueller, N.T. Can maternal-child microbial seeding interventions improve the health of infants delivered by Cesarean section? Cell Host Microbe 30, 607–611 (2022).

34. LaPoint, P., et al. Can Vaginal Seeding at Birth Improve Health Outcomes of Cesarean Section-Delivered Infants? A Scoping Review. Microorganisms 13(2025).

35. Song, S.J., et al. Naturalization of the microbiota developmental trajectory of Cesarean-born neonates after vaginal seeding. Med 2, 951–964 e955 (2021).

36. Hourigan, S.K., Mueller, N.T. & Dominguez-Bello, M.G. Can Vaginal Seeding Improve Health Outcomes of Infants Born by Cesarean Delivery? JAMA Pediatr 179, 361–362 (2025).

37. Dominguez-Bello, M.G., et al. Partial restoration of the microbiota of cesarean-born infants via vaginal microbial transfer. Nat Med 22, 250–253 (2016).

38. Mueller, N.T., et al. Maternal Bacterial Engraftment in Multiple Body Sites of Cesarean Section Born Neonates after Vaginal Seeding-a Randomized Controlled Trial. mBio 14, e0049123 (2023).

39. Zhou, L., et al. Eaects of vaginal microbiota transfer on the neurodevelopment and microbiome of cesarean-born infants: A blinded randomized controlled trial. Cell Host Microbe 31, 1232–1247 e1235 (2023).

40. Namasivayam, S., et al. Fecal transplant from vaginally seeded infants decreases intraabdominal adiposity in mice. Gut Microbes 16, 2353394 (2024).

41. Angolile, C.M., Max, B.L., Mushemba, J. & Mashauri, H.L. Global increased cesarean section rates and public health implications: A call to action. Health Sci Rep 6, e1274 (2023).

42. Stephenson, J. Rate of First-time Cesarean Deliveries on the Rise in the US. JAMA Health Forum 3, e222824 (2022).

43. Olszak, T., et al. Microbial exposure during early life has persistent eaects on natural killer T cell function. Science 336, 489–493 (2012).

44. El Aidy, S., Hooiveld, G., Tremaroli, V., Backhed, F. & Kleerebezem, M. The gut microbiota and mucosal homeostasis: colonized at birth or at adulthood, does it matter? Gut Microbes 4, 118–124 (2013).

45. Adkins, B., Leclerc, C. & Marshall-Clarke, S. Neonatal adaptive immunity comes of age. Nat Rev Immunol 4, 553–564 (2004).

46. Litvak, Y., et al. Commensal Enterobacteriaceae Protect against Salmonella Colonization through Oxygen Competition. Cell Host Microbe 25, 128–139 e125 (2019).

47. Robertson, R.C., Manges, A.R., Finlay, B.B. & Prendergast, A.J. The Human Microbiome and Child Growth - First 1000 Days and Beyond. Trends Microbiol 27, 131–147 (2019).

48. Vatanen, T., et al. Variation in Microbiome LPS Immunogenicity Contributes to Autoimmunity in Humans. Cell 165, 842–853 (2016).

49. d’Hennezel, E., Abubucker, S., Murphy, L.O. & Cullen, T.W. Total Lipopolysaccharide from the Human Gut Microbiome Silences Toll-Like Receptor Signaling. mSystems 2(2017).

50. Gewirtz, A.T., Navas, T.A., Lyons, S., Godowski, P.J. & Madara, J.L. Cutting edge: bacterial flagellin activates basolaterally expressed TLR5 to induce epithelial proinflammatory gene expression. J Immunol 167, 1882–1885 (2001).

51. Tran, H.Q., Ley, R.E., Gewirtz, A.T. & Chassaing, B. Flagellin-elicited adaptive immunity suppresses flagellated microbiota and vaccinates against chronic inflammatory diseases. Nat Commun 10, 5650 (2019).

52. Gollwitzer, E.S. & Marsland, B.J. Impact of Early-Life Exposures on Immune Maturation and Susceptibility to Disease. Trends Immunol 36, 684–696 (2015).

53. Nakamura, A., et al. Symbiotic polyamine metabolism regulates epithelial proliferation and macrophage diaerentiation in the colon. Nat Commun 12, 2105 (2021).

54. Chellappa, K., et al. NAD precursors cycle between host tissues and the gut microbiome. Cell Metab 34, 1947–1959 e1945 (2022).

55. Abdullah, N., Defaye, M. & Altier, C. Neural control of gut homeostasis. Am J Physiol Gastrointest Liver Physiol 319, G718–G732 (2020).

56. Yazji, I., et al. Endothelial TLR4 activation impairs intestinal microcirculatory perfusion in necrotizing enterocolitis via eNOS-NO-nitrite signaling. Proc Natl Acad Sci U S A 110, 9451–9456 (2013).

57. Guo, X., et al. Polyamines are necessary for synthesis and stability of occludin protein in intestinal epithelial cells. Am J Physiol Gastrointest Liver Physiol 288, G1159–1169 (2005).

58. Guo, X., et al. Regulation of adherens junctions and epithelial paracellular permeability: a novel function for polyamines. Am J Physiol Cell Physiol 285, C1174–1187 (2003).

59. Bansal, T., Alaniz, R.C., Wood, T.K. & Jayaraman, A. The bacterial signal indole increases epithelial-cell tight-junction resistance and attenuates indicators of inflammation. Proc Natl Acad Sci U S A 107, 228–233 (2010).

60. Shimada, Y., et al. Commensal bacteria-dependent indole production enhances epithelial barrier function in the colon. PLoS One 8, e80604 (2013).

61. Meng, D., et al. Indole-3-lactic acid, a metabolite of tryptophan, secreted by Bifidobacterium longum subspecies infantis is anti-inflammatory in the immature intestine. Pediatr Res 88, 209–217 (2020).

62. Mezrich, J.D., et al. An interaction between kynurenine and the aryl hydrocarbon receptor can generate regulatory T cells. J Immunol 185, 3190–3198 (2010).

63. Ho, S.W., El-Nezami, H. & Shah, N.P. The protective eaects of enriched citrulline fermented milk with Lactobacillus helveticus on the intestinal epithelium integrity against Escherichia coli infection. Sci Rep 10, 499 (2020).

64. Healy, D.B., Ryan, C.A., Ross, R.P., Stanton, C. & Dempsey, E.M. Clinical implications of preterm infant gut microbiome development. Nat Microbiol 7, 22–33 (2022).

65. Kozich, J.J., Westcott, S.L., Baxter, N.T., Highlander, S.K. & Schloss, P.D. Development of a dual-index sequencing strategy and curation pipeline for analyzing amplicon sequence data on the MiSeq Illumina sequencing platform. Appl Environ Microbiol 79, 5112–5120 (2013).

66. Bolyen, E., et al. Author Correction: Reproducible, interactive, scalable and extensible microbiome data science using QIIME 2. Nat Biotechnol 37, 1091 (2019).

67. Callahan, B.J., et al. DADA2: High-resolution sample inference from Illumina amplicon data. Nat Methods 13, 581–583 (2016).

68. McMurdie, P.J. & Holmes, S. phyloseq: an R package for reproducible interactive analysis and graphics of microbiome census data. PLoS One 8, e61217 (2013).

69. Quast, C., et al. The SILVA ribosomal RNA gene database project: improved data processing and web-based tools. Nucleic Acids Res 41, D590–596 (2013).

70. McCloskey, D., Gangoiti, J.A., Palsson, B.O. & Feist, A.M. A pH and solvent optimized reverse-phase ion-paring-LC-MS/MS method that leverages multiple scan-types for targeted absolute quantification of intracellular metabolites. Metabolomics 11, 1338–1350 (2015).

71. Cao, G., et al. Large-scale targeted metabolomics method for metabolite profiling of human samples. Anal Chim Acta 1125, 144–151 (2020).

